# Bridging field strengths: fine-tuned deep learning models for 7T MRI white matter lesion segmentation in multiple sclerosis

**DOI:** 10.64898/2026.09.03.26362180

**Authors:** Alessandro Pasquale De Rosa, Haz-Edine Assemlal, Dumitru Fetco, David Araujo, Matthew K. Schindler, Erin S. Beck, Rohit Bakshi, Daniel S. Reich, Daniel M. Harrison, Fabrizio Esposito, David A. Rudko, Douglas L. Arnold, Sridar Narayanan

## Abstract

Multiple sclerosis (MS) white matter lesion (WML) automated segmentation on ultra-high-field 7T MRI remains challenging due to the domain shift from lower-field acquisitions, limited annotated data, and specific imaging artifacts. Fine-tuning is an effective and practical strategy for adapting deep learning WML segmentation algorithms to 7T MRI, even with limited annotated data. This study evaluates fine-tuning as a domain adaptation strategy to leverage a pre-trained deep learning model for automated WML segmentation on 7T MRI. We fine-tuned a U-Net-based model, originally trained on approximately 35,000 heterogeneous lower-field (1T, 1.5T, 3T) multi-contrast MRI scans of people with MS for T2-hyperintense WML (T2-WML) segmentation, using a 7T dataset. Multiple approaches were evaluated, including standard fine-tuning on 3D FLAIR images, low-rank adaptation (LoRA) and training from scratch (nnU-Net). Models were evaluated on an external multi-center 7T test dataset. Additionally, a separate model was fine-tuned for T1-hypointense WML (T1-WML) segmentation on 7T MP2RAGE images. The original model showed substantial performance degradation on 7T data compared to 3T (Dice score decreased from 0.69 to 0.31), confirming the need for domain adaptation. Fine-tuning markedly improved T2-WML segmentation, with the fine-tuned model achieving a median Dice score of 0.57. Lesion-wise sensitivity and F1-score were 0.78 and 0.75, respectively, and the lesion volume agreement with manual segmentation was 0.93. For T1-hypointense WML, fine-tuning showed lower lesion detection performance compared to the T2-hyperintense WML segmentation (sensitivity and F1 of 0.58 and 0.50, respectively); however, incorporating multi-center data into the training set substantially reduced false positives by 23% and improved lesion detection by 14%. Multi-center fine-tuning further improved performance, particularly for the more challenging task of T1-WML segmentation. The models presented here may be included in MS research workflows to facilitate multi-center collaborations with 7T MRI.

## Introduction

Ultra-high field (UHF) MRI, such as 7 Tesla (T), has emerged as a powerful tool for advancing our understanding of multiple sclerosis (MS) pathophysiology (Louapre et al., 2020). Compared to conventional field strengths, such as 1.5T or 3T, UHF MRI allows superior spatial resolution and improved signal- and contrast-to-noise ratios (Inglese et al., 2018). These enhanced imaging characteristics enable the detection of subtle pathological changes in both white and gray matter, which may be missed at lower field strengths. Previous studies with 7T MRI on people with MS (pwMS) demonstrated a significant increase in the detectability of white matter lesions (WML), as well as a better visualization of both focal and diffuse pathology within the cortical gray matter (GM) (Kolb et al., 2021; Kollia et al., 2009; Madsen et al., 2021; Maranzano et al., 2019).

Despite the advantages of UHF MRI, the transition to 7T imaging poses new technical challenges, particularly for image acquisition and processing (Balchandani and Naidich, 2015). One major issue is the increased inhomogeneity of both the static magnetic field (B _0_) and the radiofrequency transmit field (B_1_), which are more pronounced at 7T compared to lower field strengths and can lead to substantial signal intensity variations across the image (Platt et al., 2021; Seiger et al., 2015; Stockmann and Wald, 2018). Such non-uniformities affect image quality, making the direct translation of processing and segmentation algorithms developed for lower field strengths unreliable (Cramer et al., 2025), and potentially hindering accurate automated lesion detection and quantitative assessment. Therefore, the development of reliable WML segmentation algorithms specifically tailored for 7T MRI is crucial. To date, few works have attempted to design automated algorithms for MS lesions on 7T images. Fartaria and colleagues developed an automated segmentation method for both WML and cortical lesions using a single image contrast derived from the magnetization prepared 2 rapid acquisition gradient echoes (MP2RAGE) sequence (Fartaria et al., 2019). La Rosa and colleagues trained an artificial neural network to automatically segment cortical lesions on high-resolution 7T MP2RAGE images (La Rosa et al., 2022). Manual segmentation remains the current gold standard but is extremely time-consuming and labor-intensive, particularly at 7T, where the increased spatial resolution results in a significantly higher number of voxels and slices to analyze, introducing potential inter-rater variability.

Another significant challenge is the limited availability of large, annotated multi-center datasets acquired at 7T. The scarcity of such datasets makes it difficult to develop and train deep learning (DL) models from scratch for automated lesion segmentation. Moreover, the variability in acquisition protocols and scanner hardware across sites further complicates the generalizability of DL models trained on single-center or small-scale datasets. As a result, while 7T MRI holds great promise for clinical and research applications in MS, exploiting its full potential requires alternative strategies to overcome the challenges associated with data scarcity and imaging artifacts.

One appealing approach to address the limitation of small, annotated datasets is fine-tuning. This technique involves using weights from a model that has been pre-trained on a large dataset from a related domain and then tuning it on a smaller dataset (Chen et al., 2022). In a previous work, Donnay and colleagues applied transfer learning to develop a pseudo-label assisted pipeline to automatically segment brain lesions from a single 7T MP2RAGE MRI scan. To overcome the lack of manually labeled data at 7T, the authors generated pseudo-labels on 3T data to pre-train a model; these pseudo-labels, after manual editing, were then used to fine-tune their model on 7T data (Donnay et al., 2023). To date, efforts for automated methods at 7T have all relied on the use of the T1-weighted (T1-w) MP2RAGE sequence. The fluid attenuated inversion recovery (FLAIR) sequence, which is the preferred contrast for manually and automatically identifying WML at lower field strengths, is challenging to acquire at 7T due to strong field inhomogeneities and high energy deposition, and, as such, has great quality variability across scanners (Zwanenburg et al., 2010). However, since WML pathology is more clearly visible on T2-weighted (T2-w) contrast images and relying only on MP2RAGE yields a significantly lower lesion load estimation (Zurawski et al., 2019), reliable models that work on 7T FLAIR are needed, in addition to models that work on MP2RAGE, the primary image contrast at 7T, when no FLAIR or T2w imaging is available.

In this work, we present a fine-tuned DL model for WML segmentation on 7T images that was initially pre-trained on a large, multi-center dataset consisting of thousands of MRI scans acquired at 1T, 1.5T, and 3T. We separately trained and tested models to automatically segment T2-hyperintense and T1-hypointense WML. Performance of the models was evaluated on an unseen external dataset including multi-center 7T MRI data.

## Methods

### Datasets

A large pooled and private dataset at NeuroRx, a Clario Company (Montréal, Canada), now part of Thermo Fisher Scientific, was used to train the original model for T2-WML segmentation, developed by co-author HEA, for internal use. This dataset included approximately 35,000 subjects from clinical trials acquired from 1,980 scanners with varying magnetic field strengths: 1T (0.25%), 1.5T (66%), 3T (33.75%). The conventional clinical trial protocol included 3D T1-w spoiled gradient recalled echo, 2D T2-w and proton density turbo-spin echo, and 2D T2-w FLAIR images, predominately acquired with a 1 x 1 mm^2^ in-plane resolution and 3 mm slice thickness. Only the model generated from this lower-field dataset was made available for the present research; no MRI data were provided.

Data from 28 pwMS with a 7T MRI exam were retrospectively included from two cohorts studied at the Montreal Neurological Institute-Hospital (MNI) to fine-tune the original model. Dataset 1 included 14 pwMS (mean age: 52.1 ± 8.7, 86% females), with EDSS ranging from 0 to 5.5 (median 3.0). Dataset 2 included 14 pwMS (mean age: 56.0 ± 9.7, 86% females), with EDSS ranging from 0 to 6.5 (median 2.5). An additional independent 3T dataset with a similar MRI protocol to that used at 7T was used during fine-tuning to bridge the gap between the original training data and the fine-tuning data. This dataset included 23 pwMS (mean age: 49.3 ± 13.1, 78% females), with EDSS ranging from 0 to 7.0 (median 3.5).

Finally, an external test set to evaluate the fine-tuned model was created using data extracted from the North American Imaging in MS (NAIMS) Pooled 7T common data repository (Harrison et al., 2024). This validation set included data from 30 pwMS (mean age: 42.0 ± 12.3, 56.7% females), with EDSS ranging from 0 to 6.5 (median 2.0), randomly selected from three contributing NAIMS centers (10 pwMS from each site), namely the University of Maryland, Baltimore (UMB), University of Pennsylvania (UPenn), and Brigham and Women’s Hospital (BWH) at Harvard University. Studies performed at each contributing site were approved and overseen by local institutional review boards (IRB) and participants signed informed consent, including permission for sharing of anonymized or limited data set versions of their images/data.

### MRI acquisition

All scans used for fine-tuning were acquired at the McConnell Brain Imaging Centre of the MNI (McGill University, Montréal, Canada). 7T images were acquired on a Siemens Terra scanner. The 7T MRI acquisition protocol for dataset 1 included MP2RAGE (TR/TE/TI1/TI2 = 4300/2.31/1000/3200 ms, flip angle = 4°, 0.7 mm isotropic resolution) and 3D FLAIR (TR/TE/TI = 9000/301/2250 ms, 0.7 mm isotropic resolution), acquired in single-transmit (sTx) mode. The 7T MRI acquisition protocol for dataset 2 included MP2RAGE (TR/TE/TI1/TI2 = 6000/2.74/800/2700 ms, flip angle 1 = 4°, flip angle 2 = 5°, 0.7 mm isotropic resolution) and 3D FLAIR (TR/TE/TI = 9000/301/2500 ms, 0.7 mm isotropic resolution), acquired in parallel-transmit (pTx) mode. Additional 3T images were acquired with a similar protocol on a Siemens Prisma scanner, which included MP2RAGE (TR/TE/TI1/TI2 = 5000/2.76/940/2830 ms, 1 mm isotropic resolution) and 3D FLAIR (TR/TE/TI = 6000/356/1800 ms, 1 mm isotropic resolution). 7T MRI acquisition protocols for the NAIMS external test dataset, which also included MP2RAGE and 3D FLAIR sequences, have been described in the associated publication (Harrison et al., 2024).

### Image preprocessing

MP2RAGE scans were pre-processed to obtain denoised, unified T1-w images and quantitative T1-map images (Marques et al., 2010). All the images used for fine-tuning were co-registered to the ICBM152 standard template, with voxel resolution of 1x1x1 mm and using tricubic interpolation (Fonov et al., 2011). The NAIMS images were linearly transformed to the ICBM152 template and then resampled to an isotropic 0.7 mm resolution (Harrison et al., 2024).

### Manual lesion segmentation

The original model was trained on a large, pooled clinical trial dataset, where T2-WML labels (with 1x1x3 mm voxel resolution) were initially generated using automated methods and then manually corrected by expert MRI readers (Elliott, 2016; Francis, 2004). The same automated methods and manual correction procedures were applied to the additional 3T dataset. The original model was trained on a large, pooled clinical trial dataset, where T2-WML labels (with 1x1x3 mm voxel resolution) were initially generated using automated methods and then manually corrected by expert MRI readers (Elliott, 2016; Francis, 2004). The same automated methods and manual correction procedures were applied to the additional 3T dataset. For the 7T training datasets, T2-WML labels were segmented fully manually with a minimum size of 3 mm^3^ on FLAIR scans in standard space, by one expert reader (DF). For all 7T datasets, manual masks were segmented using the FLAIR contrast as the primary image, irrespective of their visibility on MP2RAGE, and thus the resulting labels represented T2-hyperintense lesions. Additional labels representing T1-hypointense lesions were generated by applying an intensity thresholding algorithm to the MP2RAGE T1-w images within the co-registered T2-hyperintense lesion masks, followed by manual refinement on the denoised T1-w image. Lesions for the NAIMS dataset were manually annotated by two expert readers (DA and DF), who each performed an independent initial segmentation. Individual manual masks were then reviewed by both raters to reach a consensus on ambiguous lesions, and a final mask for each subject was created by the intersection of the two individual masks. Inter-rater variability was assessed on the initial segmentations by comparing total lesion number and lesion volumes, while the agreement between raters was measured with the concordance correlation coefficient (CCC).

### Original model

The original model is based on a conditional dynamic U-Net (Isensee et al., 2019). The model architecture was adapted from the 3D U-Net architecture, which is characterized by a symmetric U-shaped structure divided into encoder and decoder (Figure 1) (Çiçek et al., 2016). The encoder, or contracting path, transforms the input volume into a lower dimensional space with an increasing number of features using modular convolutional blocks. Each block consists of two smaller blocks of transformations: the first reduces the spatial dimensions of the input feature map by a factor of two, using 3x3x3 convolutional kernels and 2x2x2 stride, while in the second conditional instance normalization for cohort adaptation and Leaky ReLU activation with a negative slope of 0.01 are applied (Nichyporuk et al., 2022). The feature map is transformed again with the same set of operations with a 1x1x1 stride convolutional layer. The decoder, structured similarly but aiming to increase the spatial dimensions while reducing the encoder feature map, consists of blocks with three smaller blocks each. The first one uses transposed convolution with 2x2x2 kernels and 2x2x2 stride to double spatial dimensions. The upsampled feature map is then concatenated with the corresponding encoder feature map from the equivalent spatial level and processed through two identical smaller blocks using 3x3x3 convolutional kernels and 1x1x1 stride, with instance normalization and Leaky ReLU activation with negative slope of 0.01.

**Figure 1:**
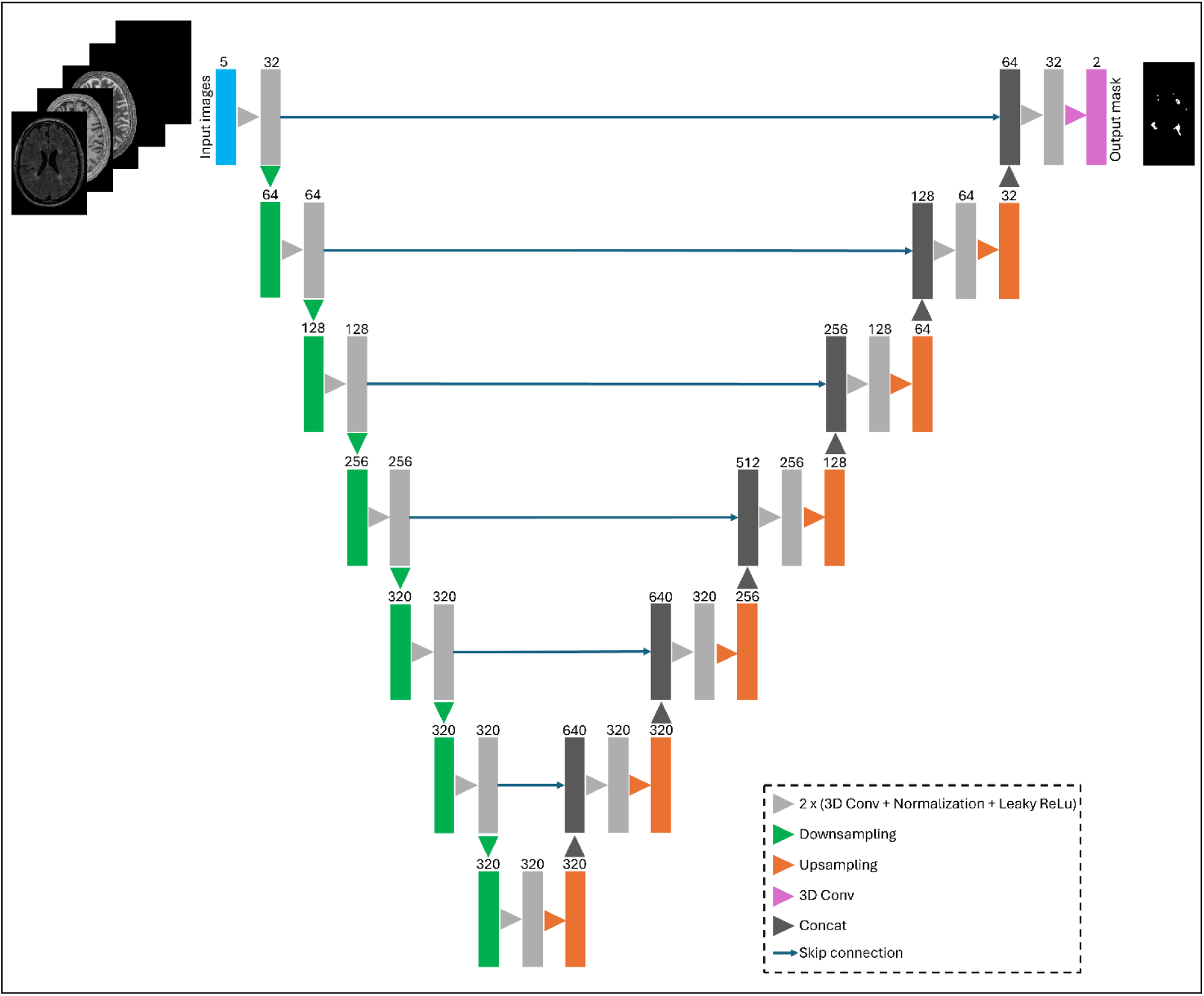
Scheme of the proposed model architecture.

During inference, 24 versions of the input volumes are generated, such that each version corresponds to one of eight possible flips along the x, y, z axes. Inference is performed for each version of the input volume, and the predictions are transformed back to the original input volume orientation by reversely applying the same flips used initially. The probabilities from all predictions are averaged, and finally each voxel is classified as background or lesion based on the class with the maximum probability.

The original model was trained using at least one (depending on their availability for each participant) of the following input scans: 3D pre-contrast T1-w, 3D post-contrast T1-w, 2D T2-weighted, 2D FLAIR and/or 2D proton density.

### Fine-tuning

Separate 3T to 7T fine-tuned models were trained for the segmentation of T2- and T1-WML. Fine-tuning was implemented by initializing the model with pre-trained weights and configurations, followed by fine-tuning the whole network on the manually segmented lesion masks from the 7T datasets (and the additional 3T dataset). The model for T2-WML segmentation used only FLAIR images as input, whereas the model for T1-WML segmentation used only MP2RAGE-derived images (T1-w and T1 maps). Fine-tuning was conducted for a maximum of 300 epochs (with early stopping after 50 epochs without improvements in validation loss), with AdamW optimizer and a maximum learning rate set to 1e-4. To adapt the decoder to the encoder, we first froze the encoder weights for 20 epochs, training only the decoder with a linearly increasing learning rate, before training the entire network. The model was trained to minimize the Tversky loss (Salehi et al., 2017). The algorithm was designed using the MONAI library v0.8.1, running with Pytorch v1.11.0 (Paszke et al., 2019).

### Data augmentation

To improve the generalizability of the model and to limit the risk of overfitting, several spatial and intensity transformations were applied to the input data during fine-tuning. Spatial transformations included random rotations (up to 0.4 radians), flipping along all three axes, zooming in, affine transformations, and foreground cropping and padding. Intensity transformations included random scaling (factor = 0.1) and shifting (offset = 0.1), contrast adjustments (gamma = [0.5, 1.5]), Rician noise addition, and simulated bias-field. Each transformation was applied with a 0.2 probability per training batch.

### Evaluation

During fine-tuning, the participants were randomly partitioned into training (70%) and validation (30%) sets. The validation set was used to assess the loss at the end of each training epoch. The final model was evaluated on the external test set. Common performance metrics were considered for the evaluation: Dice-Sørensen score (DSC) to quantify the spatial overlap between the automatic and the manual lesion masks; normalized surface Dice (NSD) to estimate which fraction of the segmented lesion boundary is correctly predicted within a tolerance τ; lesion true positive rate (LTPR) to measure the proportion of true lesions that are successfully detected, lesion false discovery rate (LFDR) to measure the proportion of segmented lesions that do not correspond to any true lesion; lesion F1-score (LF1) as summary measure of lesion-detection precision and lesion-detection recall; CCC between manually-derived and segmented volume estimates. The tolerance τ for NSD was chosen by estimating the average surface distance (ASD) between the individual manual masks of two raters on the 7T NAIMS dataset to capture the natural human variability in manually delineating the lesions. Lesion detection performance was also evaluated via TPR-FDR curves and area under curve (AUC). Finally, we compared original and fine-tuned models at different minimum lesion volumes (1, 3, 6, 9, and 15 mm^3^). Ablation studies were also conducted to evaluate the performance of different input combinations and across different lesion size thresholds.

All experiments were conducted on a Linux-based workstation equipped with a NVIDIA RTX A6000 GPU (48 GB VRAM).

### Comparison with other methods

We evaluated the performance of T2-WML segmentation on 7T data from two recently published state-of-the-art models, both trained and evaluated on 3T or lower field strength data:

• FLAMeS, a deep learning-based model trained on 668 FLAIR 1.5T and 3T scans from pwMS (Dereskewicz et al., 2025).

• MindGlide, a deep learning model trained on 4,247 brain MRI scans (1.5T and 3T) from 2,934 pwMS across 592 scanners (Goebl et al., 2025).

For comparison, we also evaluated models trained from scratch and a different fine-tuning approach:

• nnU-Net models were trained from scratch exclusively on the 7T data for 200 epochs using the nnunetv2 package (https://github.com/MIC-DKFZ/nnUNet). The nnU-Net framework extracts dataset properties such as image size, voxel spatial information, and category distribution, to automatically adjust hyperparameters, guiding the construction and data manipulation of the neural network (Isensee et al., 2021).

• Low-rank adaptation (LoRA) is a fine-tuning technique that freezes the pre-trained model weights and injects trainable rank decomposition matrices (Hu et al., 2021). This approach, originally designed for large language models, significantly reduces the number of trainable parameters and memory usage, making the fine-tuning process more efficient. Similarly to full fine-tuning, LoRA was applied to the original model using 7T data and the additional 3T dataset.

The Wilcoxon signed-rank test was used for pairwise comparisons. Differences were considered significant for p-value < 0.05.

## Results

### Manual segmentation

Figure 2 shows lesion count and lesion load distributions across sites for T1- and T2-WML lesions. In the NAIMS dataset, T2 lesion counts showed high concordance between raters (CCC = 0.97), while T2 lesion volumes showed lower agreement (CCC = 0.85), with Rater 1 consistently segmenting larger lesion volumes on average (p < 0.001) (Supplementary Table 1). The median ASD between the two individual masks was 0.71 mm; this value (≈ 1 voxel at 0.7 mm isotropic resolution) was set as boundary tolerance τ for NSD computation.

**Figure 2:**
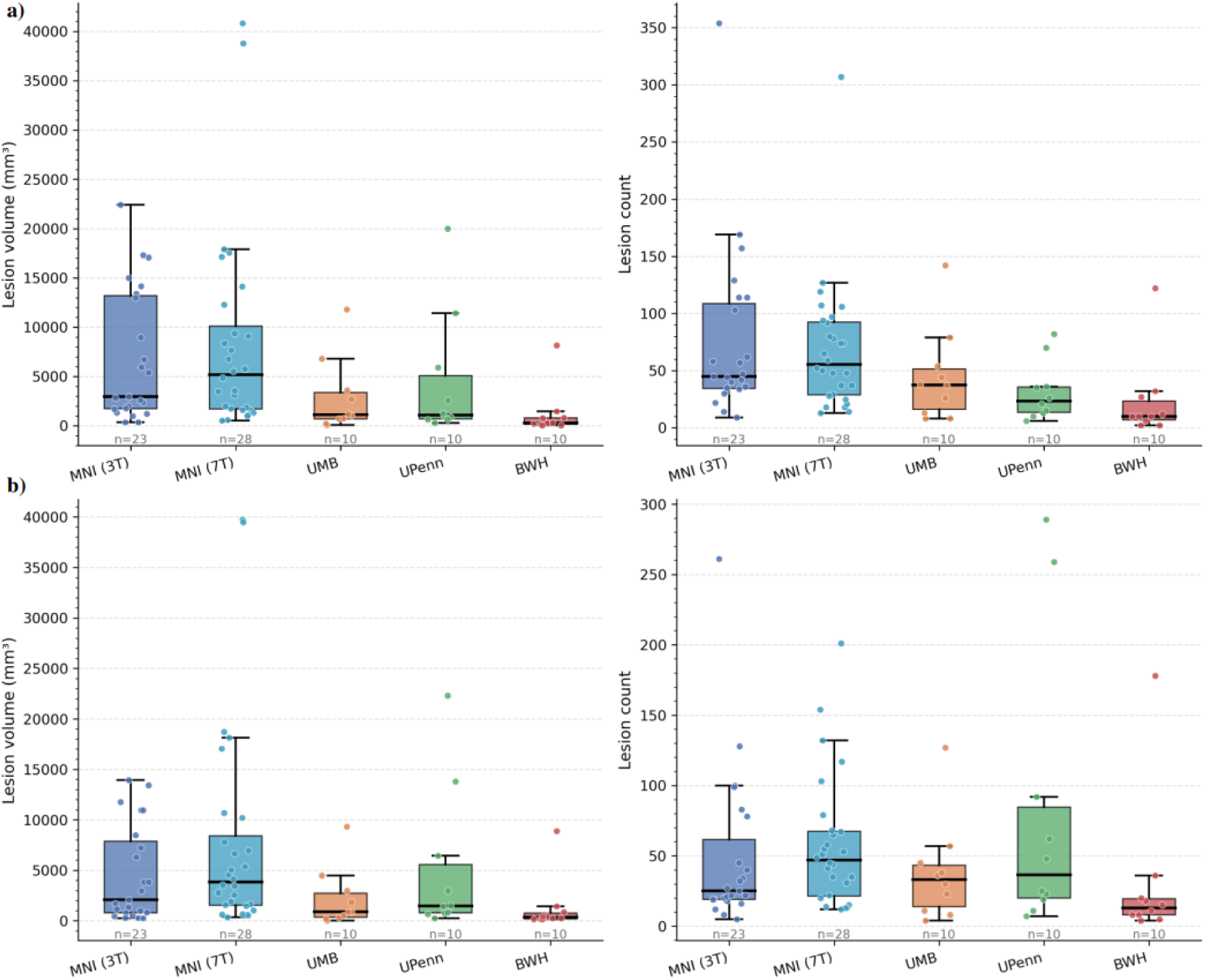
Lesion load and lesion count distributions across sites from manual T2-WML segmentation (a) and from manual T1-WML segmentation (b). Abbreviations: Montréal Neurological Institute (MNI), University of Maryland, Baltimore (UMB), University of Pennsylvania (UPenn), Brigham and Women’s Hospital (BWH).

### Original model performance on 3T dataset

The original model was initially trained to work on 1.5T and 3T MRI data for segmenting T2-WML. We evaluated its performance on the additional 3T MNI dataset. Results are reported in Supplementary Table 2.

### Fine-tuned model performance on 7T dataset

Figure 3 shows the automatically segmented lesion masks for a representative participant from each site of the NAIMS dataset, alongside the corresponding manual segmentations and input images used with the model. Model performance was assessed by comparing the segmented lesions masks to the ground truth references. Median evaluation metrics for the original model, fine-tuned model, nnU-Net model, and LoRA model on the external test dataset are summarized in Table 1.

**Figure 3:**
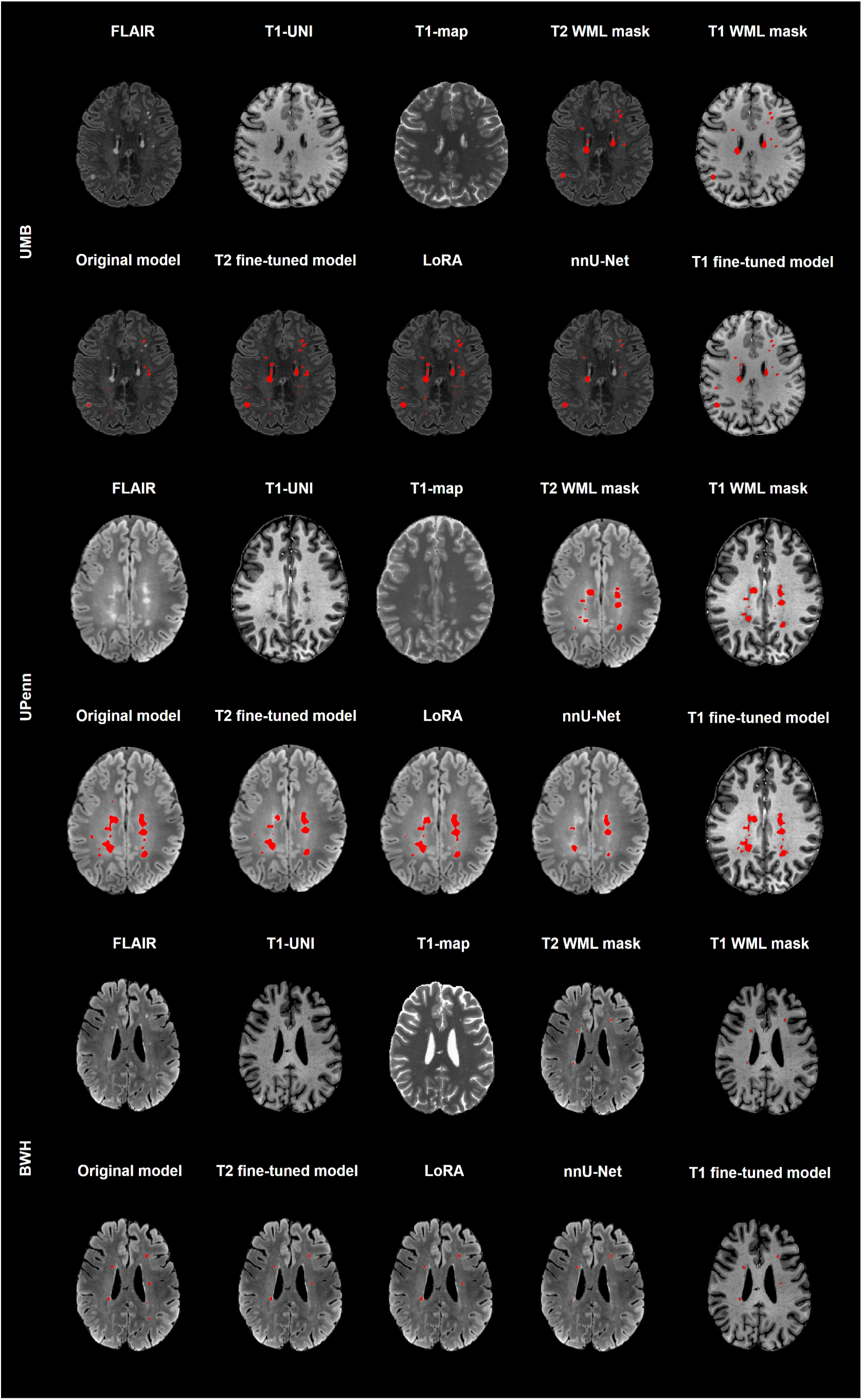
Visual comparison between manually segmented lesion masks and automatically segmented lesion masks from each evaluated method. One participant for each site of the NAIMS test dataset is showed. Abbreviations: University of Maryland, Baltimore (UMB); University of Pennsylvania (UPenn); Brigham and Women’s Hospital (BWH).

**Table 1:** Median evaluation metrics across test subjects for each model. Best values are in bold. Abbreviations: Dice-Sørensen score (DSC); normalized surface Dice (NSD); lesion true positive rate (LTPR); lesion false discovery rate (LFDR); lesion F-score (LF1).

|  | T2-hyperintense lesion segmentation |  |  |  |  |
| --- | --- | --- | --- | --- | --- |
|  | DSC | NSD | LTPR | LFDR | LF1 |
| **Original model** | 0.31 | 0.39 | 0.66 | 0.55 | 0.52 |
| **Fine-tuned model** | **0.57** | **0.69** | 0.78 | 0.27 | **0.75** |
| **nnU-Net** | 0.52 | 0.59 | 0.65 | **0.21** | 0.71 |
| **LoRA** | 0.56 | 0.65 | **0.82** | 0.36 | 0.70 |
|  | T1-hypointense lesion segmentation |  |  |  |  |
|  | DSC | NSD | LTPR | LFDR | LF1 |
| **Fine-tuned model** | **0.54** | 0.60 | **0.58** | 0.53 | 0.50 |
| **nnU-Net** | 0.47 | 0.55 | 0.41 | **0.23** | 0.52 |
| **LoRA** | **0.54** | **0.66** | 0.49 | 0.48 | **0.56** |

The original model showed significant performance degradation on 7T data, with DSC and LF1 dropping by 38% and 23%, respectively. Segmented lesion masks from FLAMes and MindGlide resulted in similar or lower performance metrics (Supplementary Table 3). The fine-tuned model significantly outperformed the original model for the T2-WML task in terms of median DSC (0.57 vs. 0.31, p < 0.001), NSD (0.69 vs. 0.39, p < 0.001), LTPR (0.78 vs. 0.66, p < 0.01), LFDR (0.27 vs. 0.55, p < 0.001), and LF1 (0.75 vs. 0.52, p < 0.001). Similarly, both constrained fine-tuning with LoRA and the nnU-Net model trained from scratch clearly performed better than the original model. The T1-WML segmentation task showed lower performance compared to the T2-WML segmentations task, with the LoRA approach reaching the highest DSC (0.54), NSD (0.66), and LF1 (0.56) scores, and the full fine-tuned model having the best LTPR (0.58). Figure 4 shows all five performance metrics for the fine-tuned model stratified by NAIMS site and tasks.

**Figure 4:**
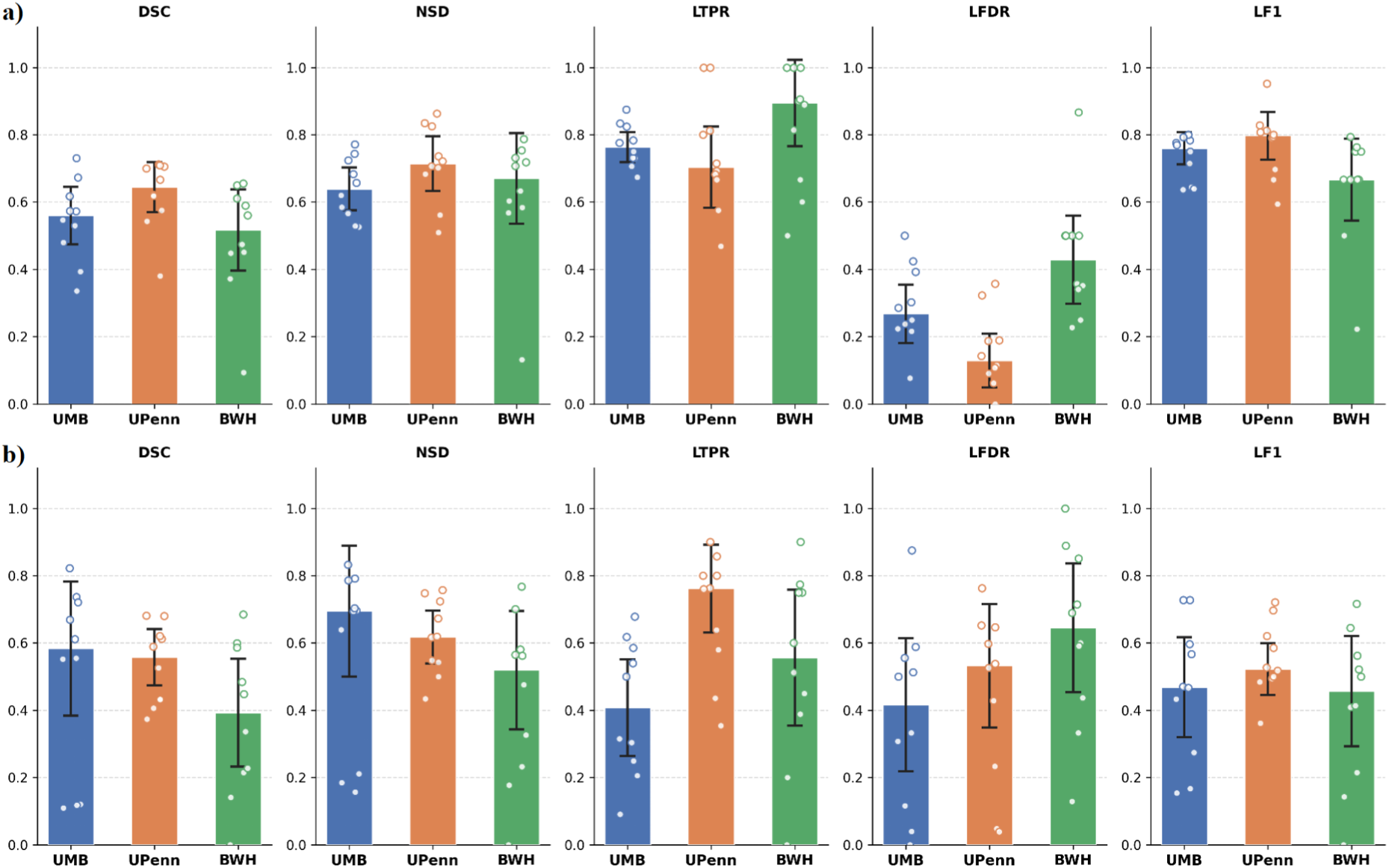
Performance metrics across acquisition sites for T2-hyperintense WML segmentation (a) and T1-hypointense WML segmentation. Bar plots show median values (± 95% confidence interval) for all segmentation metrics. Individual observations are overlaid as scatter points. Abbreviations: Dice-Sørensen score (DSC); normalized surface Dice (NSD); lesion true positive rate (LTPR); lesion false discovery rate (LFDR); lesion F-score (LF1); University of Maryland, Baltimore (UMB); University of Pennsylvania (UPenn); Brigham and Women’s Hospital (BWH).

Table 2 reports T2-WML detection rates across different lesion sizes. Lesions were categorized into three classes (small, medium, and large) based on the tertiles of their volume distribution across all participants. In the 30 cases from the NAIMS dataset, a total of 996 manually segmented T2-WML were identified (median volume [interquartile range]: 15.4 [31.6] mm^3^). The results showed that fine-tuning consistently improved lesion segmentation across all size classes, with performance increasing with lesion size.

**Table 2:** Comparison of WML segmentation performance between the original model and the fine-tuned model for different lesion size categories. Lesion-level metrics for the external NAIMS dataset are reported as median values for different ranges of lesion size. Best values are in bold. Abbreviations: lesion true positive rate (LTPR); lesion false discovery rate (LFDR); lesion F-score (LF1).

|  | **LTPR** |  | **LFDR** |  | **LF1** |  |
| --- | --- | --- | --- | --- | --- | --- |
|  | Original<br>model | Fine-tuned<br>model | Original<br>model | Fine-tuned<br>model | Original<br>model | Fine-tuned<br>model |
| **Small lesions**<br>**[1, 9.6][mm<sup>3</sup>]** | 0.38 | **0.40** | 0.79 | **0.74** | 0.29 | **0.31** |
| **Medium lesions**<br>**(9.6, 27.4][mm<sup>3</sup>]** | 0.62 | **0.77** | 0.46 | **0.24** | 0.59 | **0.77** |
| **Large lesions**<br>**(> 27.4)[mm<sup>3</sup>]** | 0.73 | **0.88** | 0.41 | **0.12** | 0.70 | **0.88** |

TPR-FDR curves (Figure 5a) showed that the fine-tuned models also had the best T2-WML detection performance (AUC = 0.77 for both full and LoRA fine-tuning). TPR-FDR curves for T1-WML segmentation (Figure 5b) showed AUC values of 0.54 for full and LoRA fine-tuned models, and 0.58 for the nnU-Net model.

**Figure 5:**
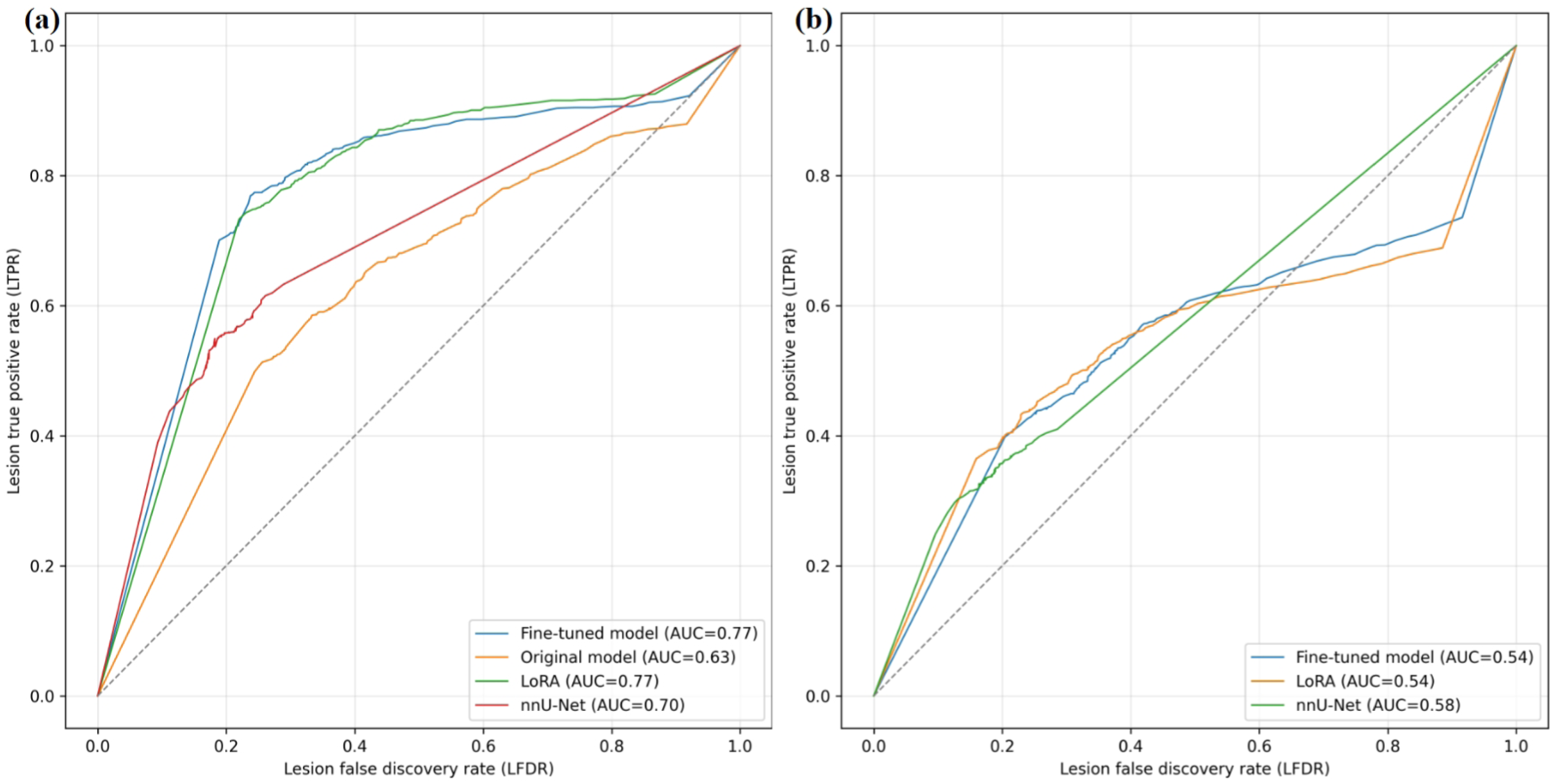
Comparison of lesion-wise TPR-FDR curves for T2-hyperintense lesion segmentation (left) and T1-hypointense lesion segmentation (right) between different lesion segmentation algorithms on the NAIMS test dataset.

Finally, the fine-tuned models showed the highest concordance with both T2 (CCC = 0.93, Figure 6) and T1 (CCC = 0.98) and T1 (CCC = 0.98, Figure 7) manually segmented lesion volumes compared to the other models.

**Figure 6:**
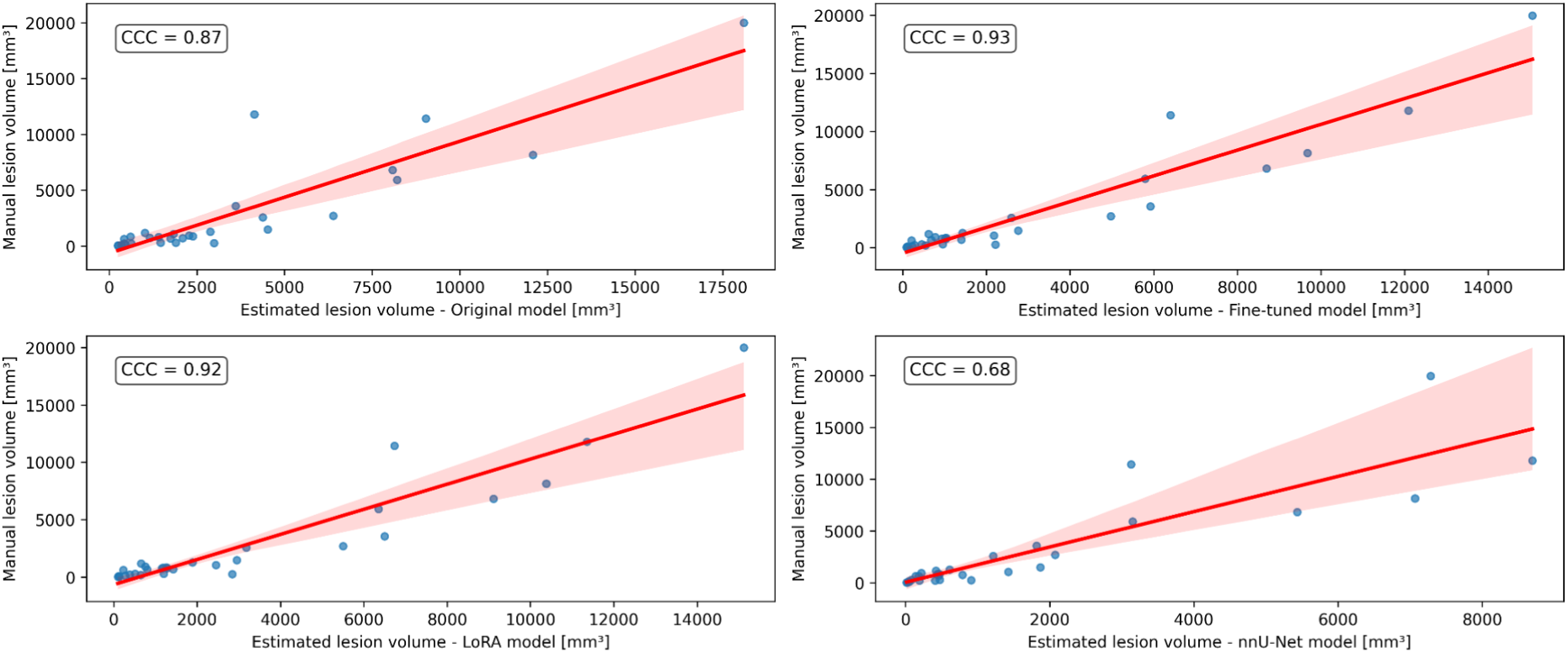
Correlations between manual T2 lesion volumes and automatically estimated T2 lesion volumes. The solid line shows the linear regression line along with a confidence interval at 95% for the original model (top-left), fine-tuned model (top-right), LoRA model (bottom-left), and nnU-Net model (bottom-right). Abbreviations: concordance correlation coefficient (CCC).

**Figure 7:**
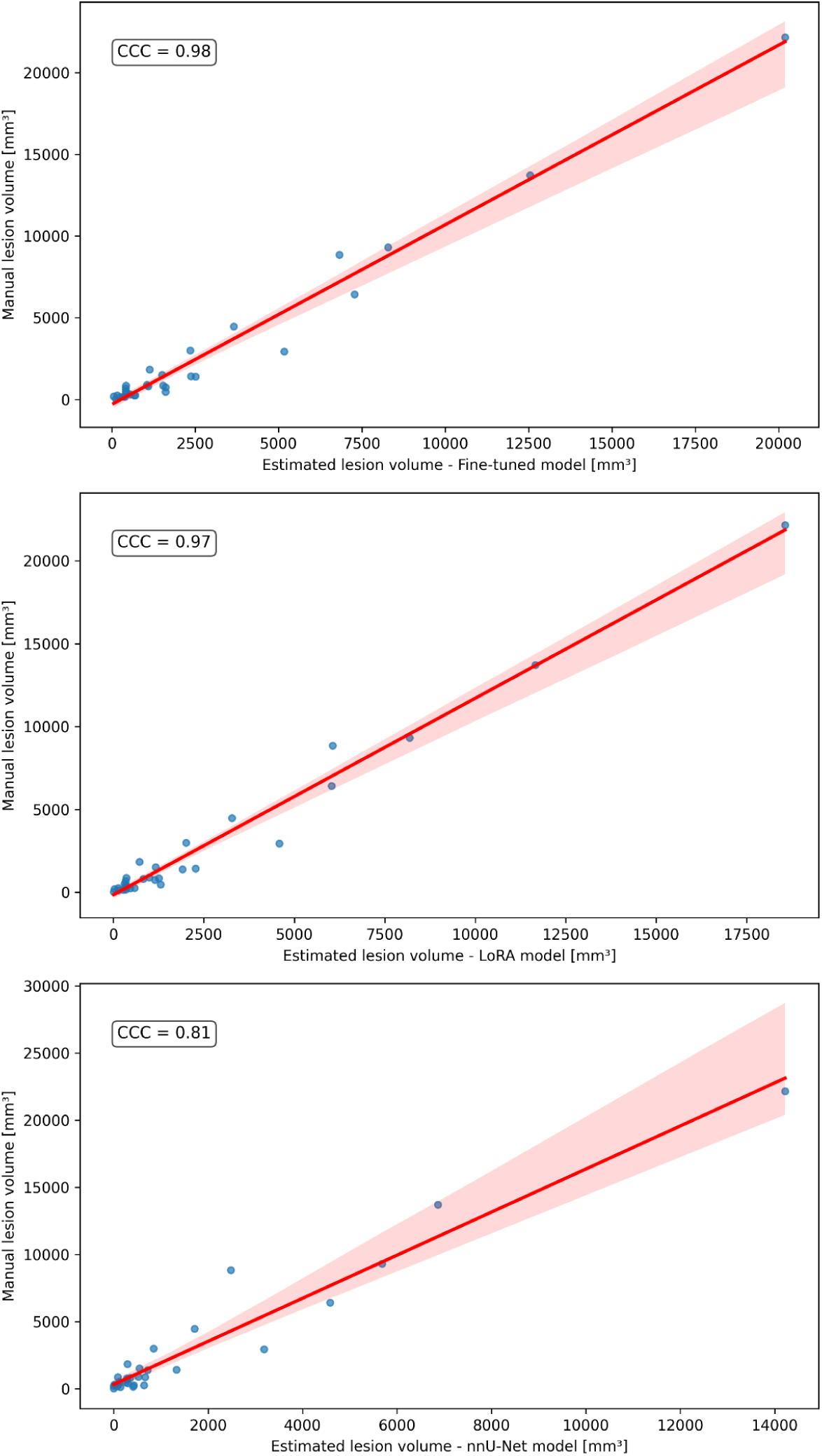
Correlations between manual T1 lesion volumes and automatically estimated T1 lesion volumes. The solid line shows the linear regression line along with a confidence interval at 95% for the fine-tuned model (top), LoRA model (middle), and nnU-Net model (bottom). Abbreviations: concordance correlation coefficient (CCC).

### Fine-tuning on multi-center dataset for T1-hypointense WML segmentation

Since the T1-WML segmentation model performed considerably worse than its T2-WML counterpart, we hypothesized that the greater difficulty of delineating lesions on MP2RAGE alone would demand a more diverse and heterogeneous training dataset. To address this issue, we fine-tuned the model by including MP2RAGE images from multi-site data into the training set. We used a stratified (by site) 5-fold cross-validation procedure to ensure that each of the 58 participants with 7T MRI was included in the test set exactly once. Multi-center training substantially reduced false positive proportion (LFDR dropping from 0.53 to 0.30, p < 0.001) and improved overall lesion detection (LF1 increasing from 0.50 to 0.64, p < 0.001) (Table 3). Subjects from the MNI site yielded the best segmentation performance. Multi-center training substantially reduced false positive proportion (LFDR dropping from 0.53 to 0.30, p < 0.001) and improved overall lesion detection (LF1 increasing from 0.50 to 0.64, p < 0.001) (Table 3). Subjects from the MNI site yielded the best segmentation performance.

**Table 3:** Median evaluation metrics on test subjects after fine-tuning the T1-WML model on MP2RAGE images from multi-center training data. Abbreviations: Montréal Neurological Institute (MNI), University of Maryland, Baltimore (UMB), University of Pennsylvania (UPenn), Brigham and Women’s Hospital (BWH), Dice-Sørensen score (DSC); normalized surface Dice (NSD); lesion true positive rate (LTPR); lesion false discovery rate (LFDR); lesion F-score (LF1).

|  | **DSC** | **NSD** | **LTPR** | **LFDR** | **LF1** |
| --- | --- | --- | --- | --- | --- |
| **UMB** | 0.48 | 0.53 | 0.35 | 0.29 | 0.45 |
| **UPenn** | 0.55 | 0.66 | 0.68 | 0.30 | 0.64 |
| **BWH** | 0.51 | 0.59 | 0.45 | 0.31 | 0.50 |
| **MNI** | 0.64 | 0.75 | 0.64 | 0.18 | 0.67 |
| **Overall** | 0.59 | 0.62 | 0.58 | 0.30 | 0.64 |

### Leave-one-site-out cross-validation

To futher assess the heterogeneity across sites, we also run a leave-one-site-out cross-validation (LOSOCV), where each of the 3 NAIMS sites was used as external testing set for both T2-WML and T1-WML segmentation. Results are reported in Table 4. For the T2-WML segmentation task, NSD ranged between 0.61 and 0.75, while overall lesion detection (LF1) between 0.65 and 0.73, showing consistent performance across sites. For the T1-WML segmentation task, larger variation was observed, with NSD ranging from 0.45 to 0.64 and LF1 between 0.38 and 0.64.

**Table 4:** Median evaluation metrics on held-out sites after leave-one-site-out cross-validation. Abbreviations: University of Maryland, Baltimore (UMB), University of Pennsylvania (UPenn), Brigham and Women’s Hospital (BWH), Dice-Sørensen score (DSC); normalized surface Dice (NSD); lesion true positive rate (LTPR); lesion false discovery rate (LFDR); lesion F-score (LF1).

|  | **DSC** | **NSD** | **LTPR** | **LFDR** | **LF1** |
| --- | --- | --- | --- | --- | --- |
| **T2-hyperintense lesion segmentation** |  |  |  |  |  |
| **UMB** | 0.56 | 0.61 | 0.63 | 0.27 | 0.65 |
| **UPenn** | 0.55 | 0.63 | 0.66 | 0.04 | 0.73 |
| **BWH** | 0.62 | 0.75 | 0.85 | 0.40 | 0.72 |
| **T1-hyperintense lesion segmentation** |  |  |  |  |  |
| **UMB** | 0.57 | 0.64 | 0.26 | 0.24 | 0.38 |
| **UPenn** | 0.43 | 0.45 | 0.76 | 0.34 | 0.64 |
| **BWH** | 0.45 | 0.54 | 0.42 | 0.36 | 0.53 |

### Ablation studies

To evaluate the contribution of each image contrast to the overall performance, we added the MP2RAGE-derived images (T1-w denoised and T1-map) to the T2-WML segmentation model and removed the T1-map from the T1-WML segmentation model. In neither case did the modification to the model inputs show a benefit (Supplementary Table 4). Supplementary Figure 1 shows the LTPR and LFDR of all models across different minimum T2 lesion volumes (i.e. 1, 3, 6, 9, and 15 mm^3^). The fine-tuned model showed a consistent performance across all lesion size cutoffs in terms of LTPR (≈ 0.80) and LFDR (≈ 0.27, from the 3 mm^3^ cutoff). LoRA showed a similar pattern for LTPR, while being less consistent for LFDR.

## Discussion

Our work aimed to show that fine-tuning can be effectively employed to adapt a pre-trained model for the automated segmentation of WML of 7T MRI data. For this purpose, we fine-tuned and evaluated a U-Net-based model originally trained for T2-WML segmentation on a large, heterogeneous dataset consisting of roughly 35,000 lower-field MRI scans (at 1T, 1.5T, and 3T) of pwMS. We analyzed and compared different models, using multi-contrast inputs and approaches, based on their performance on an external, multi-center 7T test dataset. By fine-tuning this model with a relatively small 7T dataset and an intermediate 3T dataset with harmonized acquisition protocol, we aimed to improve T2-WML segmentation accuracy on 7T FLAIR images. We further evaluated the efficacy of fine-tuning as a domain adaptation strategy, re-training the original T2-w/FLAIR-based model for T1-WML segmentation on 7T MP2RAGE images. This was clinically motivated by the frequent absence of FLAIR sequences in current 7T MRI acquisition protocols, where MP2RAGE is often the only available contrast for WML detection.

### T2-WML segmentation performance

The original model showed robust T2-WML segmentation performance on the unseen 3T dataset (Supplementary Table 2), achieving performance metrics comparable to those reported for recent state-of-the-art models (Cerri et al., 2021; Dereskewicz et al., 2025). Conversely, its performance was substantially reduced when applied to the 7T MRI dataset (Table 1), likely reflecting the domain shift between 3T and 7T data, and thus justifying the need for an automated model tailored to 7T. Recently published state-of-the-art models (i.e. FLAMeS and MindGlide) also performed poorly on 7T data, further confirming that models pretrained on lower field strength data struggle on UHF data with performance degradation.

Compared to the original model, the fine-tuned model substantially improved voxel-level metrics such as DSC (0.57, p < 0.001) and NSD (0.69, p < 0.001) indicating better spatial overlap and boundary delineation of lesions. Lesion-level metrics revealed a significantly higher LTPR (0.78, p < 0.001) and F1 (0.75, p < 0.001) for the fine-tuned model, a critical improvement for reducing the manual correction burden. LoRA, a memory-efficient fine-tuning strategy originally developed for large language models, also showed similar improvements (Table 1), suggesting that it represents a valid alternative in medical imaging segmentation when computational resources are limited. In contrast, a model trained from scratch, based on the nnU-Net architecture, performed worse in terms of lesion detection, failing to consistently segment lesions, possibly due to underfitting; this result confirms the challenges of adequately training robust DL models from scratch with limited availability of annotated 7T data. Moreover, the fine-tuned model consistently improved lesion-level segmentation regardless of the lesion size. Although segmentation of small lesions (< 10 mm^3^) proved to be challenging (LTPR = 0.40), our model improved substantially with increasing lesion size across all participants, achieving high accuracy for medium- and large-sized lesions (LTPR of 0.77 and 0.88, respectively). Additionally, in the LOSOCV, T2-WML segmentation with the fine-tuned model showed to be consistent across sites, yielding comparable performance metrics (Table 4). Overall, the fine-tuned models showed the best segmentation and lesion detection performance, as also indicated by the TPR-FDR curves, with AUC of 0.77. Moreover, it proved to be highly consistent and performed well with different minimum lesion volume thresholds (i.e. 1, 3, 6, 9, and 15 mm^3^), with a lesion-level detection rate around 80% across all cutoffs (Supplementary Figure 1).

The challenge of FLAIR imaging at 7T is well recognized, due to increased bias field inhomogeneities and artifacts which lead to contrast and signal loss within the images, especially in the temporal lobes and cerebellum (Opheim et al., 2021). Additionally, these issues translate into greater variability of image quality across scanners and sites. However, despite these challenges, FLAIR images may still benefit from the increased signal- and contrast-to-noise ratios at UHF strengths, similarly to other sequences (Zwanenburg et al., 2010), increasing lesion detectability. Moreover, previous work on 7T showed that estimated lesion volume derived from FLAIR images is consistently higher compared to MP2RAGE images, arguably due to larger individual lesion volumes (Spini et al., 2020). Although previous attempts to develop an automated method for MS lesion segmentation on 7T data focused solely on MP2RAGE, designing an algorithm that works with FLAIR images is valuable for several applications. For example, recent works on paramagnetic rim lesions (PRL) used FLAIR images to manually segment WML before determining if an iron rim was present or not (Dal-Bianco et al., 2021; Marshall et al., 2025). For automatically segmenting PRL at 7T, T2-hyperintense WML masks may serve as initial search space for the model.

### T1-WML segmentation performance

T1-hypointense WML segmentation proved to be a more challenging task for a fine-tuning approach. This required shifting the original domain from detecting T2-hyperintensities to detecting T1-hypointensities, a more demanding adaptation given the inverted contrast and generally lower lesion conspicuity on MP2RAGE images. Indeed, even if the fine-tuned model outperformed the nnU-Net model and showed high consistency with manually-derived T1 lesion volumes (CCC = 0.98), lesion-detection performance was suboptimal (LTPR = 0.58, LFDR = 0.53, LF1 = 0.50, AUC = 0.54). Including a multi-center dataset into the fine-tuning process led to substantial gains in model performance, especially in terms of reducing the proportion of false positives (LFDR = 0.30, p < 0.001), i.e. increasing precision, and increasing the overall lesion detection (LF1 = 0.64, p < 0.001). Expectedly, subjects from the MNI site (i.e. the largest and most heterogeneous subgroup) yielded the best segmentation performance (Table 3), suggesting that the model benefits directly from increased sample size and acquisition diversity. However, the LOSOCV showed heterogenous results across sites (Table 4), especially if compared to the T2-WML segmentation task. Taken together, these results demonstrate that multi-center fine-tuning is a viable and effective strategy for T1-hypointense WML segmentation, and that our multi-center fine-tuned model may already constitute a valid benchmark for future developments, with performance expected to improve further as larger and more diverse multi-center 7T datasets become available.

### Perspectives and conclusions

Previous 7T MRI studies in MS research have been mainly conducted on single-center datasets with small sample sizes (typically fewer than 50 participants). With the growing availability of 7T MRI scanners and the emergence of multi-center collaborations, such as NAIMS, future studies are expected to include larger and more heterogeneous populations (Harrison et al., 2024). Therefore, it is of paramount importance, for consistency and reproducibility, to design reliable and robust approaches for automated WML detection to help reduce the manual effort for lesion segmentation, which remains a time-consuming and subjective task.

Despite our efforts of pooling together a multi-center dataset, the total number of annotated 7T scans was still relatively small. Therefore, model generalizability across different hardware vendors, sequence implementations, and diverse lesion characteristics remains to be tested on larger datasets. In this context, the NAIMS cooperative is addressing the need for standardization of 7T MRI protocols in MS, which would be beneficial for improving the generalization of automated methods.

While this was out of the scope of this work, the ability to detect cortical lesions remains a significant need in MS imaging (Calabrese et al., 2010; Madsen et al., 2021). Future work could explore transfer learning strategies for leukocortical lesions, which are often small and harder to annotate consistently, but detectable even at 3T (Maranzano et al., 2019). Moreover, future research should also aim to integrate model uncertainty estimation: quantifying confidence in predictions would allow clinicians to better interpret model outputs and potentially flag uncertain regions for expert revision (Nair et al., 2020). These improvements would be beneficial for real-world deployment of automated segmentation tools for 7T MRI in clinical practice.

In conclusion, this work demonstrates that fine-tuning is currently a valid strategy to adapt DL models trained on lower-field MRI data for use in 7T MRI. Even with a relatively small number of high-quality annotated 7T scans, we successfully fine-tuned a segmentation model that outperforms the original model on unseen multi-center data. We also introduced a model trained exclusively on T1-weighted images derived from 7T MP2RAGE scans, addressing the practical scenario in which FLAIR imaging is absent from the 7T acquisition protocol. As availability of 7T MRI increases, but the lack of large, annotated datasets remains, fine-tuned models like the ones presented here may facilitate the integration of automated segmentation into multi-center research and potentially clinical workflows.

## Data Availability

All data produced in the present study are available upon reasonable request to the authors.

## Acknowledgements

Acquisition of 7T MRIs from the MNI were acquired under Canadian Institutes of Health Research FRN 84367 and PJT153005, and the United States Department of Defense, Multiple Sclerosis Research Program, Investigator-Initiated Research Award (Award No. W81XWH19-1-0486). Aggregation of 7T data included in the NAIMS-IR was made possible by NIH grant R01NS122980, while funding for the NAIMS-IR itself was provided by the Hilton Foundation Grant number 20220. This research was also supported in part by the Intramural Research Program of the National Institutes of Health (NIH). The contributions of the NIH authors are considered Works of the United States Government. The findings and conclusions presented in this paper are those of the authors and do not necessarily reflect the views of the NIH or the US Department of Health and Human Services.

---

**Supplementary Figure 1:**
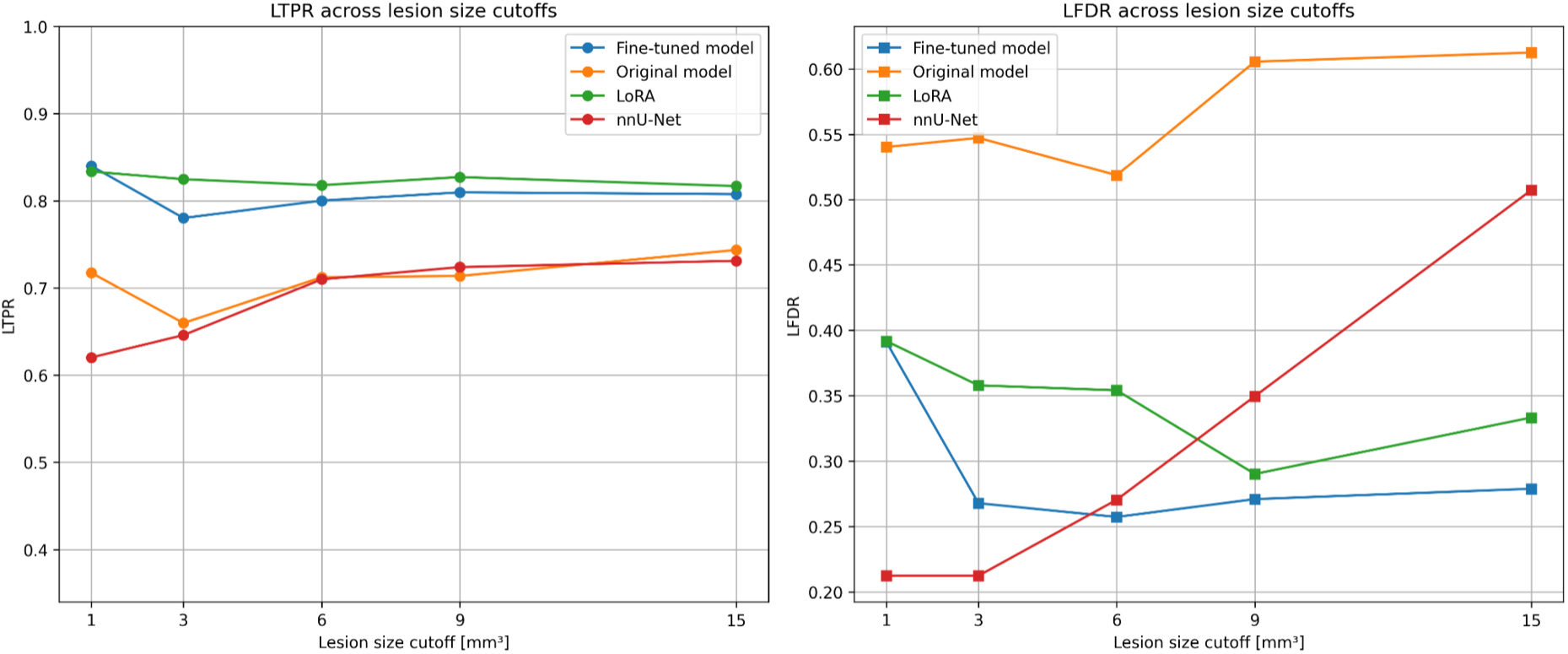
T2-hyperintense lesion detection scores for different lesion size cutoffs. Abbreviations: lesion true positive rate (LTPR); lesion false discovery rate (LFDR).

**Supplementary Table 1:** T2 lesion counts and lesion volumes determined by the two raters in the testing NAIMS dataset. Values are reported as median [interquartile range]. Significant p-values from the comparison between raters with the Wilcoxon rank-sum test are in bold. Abbreviations: concordance correlation coefficient (CCC).

|  | Rater 1 | Rater 2 | Intersection | CCC | p-value |
| --- | --- | --- | --- | --- | --- |
| **Count** | 28 [30] | 24 [28] | 24 [28] | 0.97 | 0.19 |
| **Volume**<br>[mm<sup>3</sup>] | 1725 [4692] | 857 [2390] | 847 [2378] | 0.85 | **< 0.001** |

**Supplementary Table 2:** Median evaluation metrics of the original model for T2-hyperintense WML segmentation on the 3T MNI dataset. Abbreviations: Dice-Sørensen score (DSC); lesion true positive rate (LTPR); lesion false discovery rate (LFDR); F-score (LF1).

|  | DSC | LTPR | LFDR | LF1 |
| --- | --- | --- | --- | --- |
| **Original model** | 0.69 | 0.66 | 0.11 | 0.75 |

**Supplementary Table 3:**
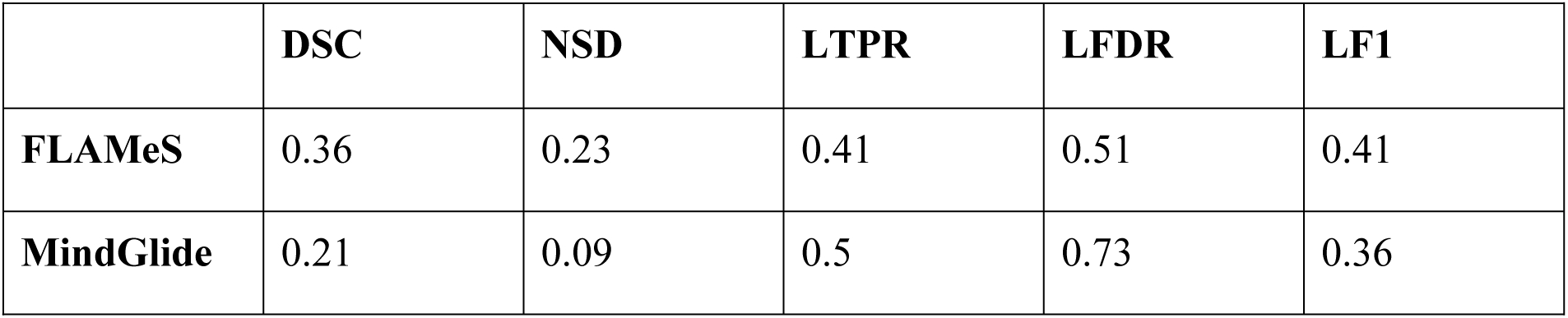
Median evaluation metrics of recently published state-of-the-art models for T2-hyperintense WML segmentation on the 7T NAIMS dataset. Abbreviations: Dice-Sørensen score (DSC); normalized surface Dice (NSD); lesion true positive rate (LTPR); lesion false discovery rate (LFDR); F-score (LF1).

|  | DSC | NSD | LTPR | LFDR | LF1 |
| --- | --- | --- | --- | --- | --- |
| **FLAMeS** | 0.36 | 0.23 | 0.41 | 0.51 | 0.41 |
| **MindGlide** | 0.21 | 0.09 | 0.5 | 0.73 | 0.36 |

**Supplementary Table 4:** Median evaluation metrics across test subjects for ablation studies, i.e. adding MP2RAGE-derived images to the T2-hyperintense lesion segmentation model and removing the T1-map from the T1-hypointense lesion segmentation model. Abbreviations: Dice-Sørensen score (DSC); normalized surface Dice (NSD); lesion true positive rate (LTPR); lesion false discovery rate (LFDR); F-score (LF1).

| **T2-hyperintense lesion segmentation** |  |  |  |  |  |
| --- | --- | --- | --- | --- | --- |
|  | DSC | NSD | LTPR | LFDR | LF1 |
| **Fine-tuned model** | 0.56 | 0.62 | 0.71 | 0.35 | 0.68 |
| **T1-hypointense lesion segmentation** |  |  |  |  |  |
|  | DSC | NSD | LTPR | LFDR | LF1 |
| **Fine-tuned model** | 0.47 | 0.55 | 0.54 | 0.52 | 0.49 |

